# Patient-level detection of grade group ≥2 prostate cancer using quantitative diffusion MRI

**DOI:** 10.1101/2021.05.24.21256461

**Authors:** Allison Y. Zhong, Leonardino A. Digma, Troy Hussain, Christine H. Feng, Christopher C. Conlin, Karen Tye, Asona J. Lui, Maren M.S. Andreassen, Ana E. Rodríguez-Soto, Roshan Karunamuni, Joshua Kuperman, Christopher J. Kane, Rebecca Rakow-Penner, Michael E. Hahn, Anders M. Dale, Tyler M. Seibert

## Abstract

**Purpose:** Multiparametric MRI (mpMRI) improves detection of clinically significant prostate cancer (csPCa), but the qualitative PI-RADS system and quantitative apparent diffusion coefficient (ADC) yield inconsistent results. An advanced Restrictrion Spectrum Imaging (RSI) model may yield a better quantitative marker for csPCa, the RSI restriction score (RSI_rs_). We evaluated RSI_rs_ for patient-level detection of csPCa.

**Materials and Methods:** Retrospective analysis of men who underwent mpMRI with RSI and prostate biopsy for suspected prostate cancer from 2017-2019. Maximum RSI_rs_ within the prostate was assessed by area under the receiver operating characteristic curve (AUC) for discriminating csPCa (grade group ≥2) from benign or grade group 1 biopsies. Performance of RSI_rs_ was compared to minimum ADC and PI-RADS v2-2.1via bootstrap confidence intervals and bootstrap difference (two-tailed α=0.05). We also tested whether the combination of PI-RADS and RSI_rs_ (PI-RADS+RSI_rs_) was superior to PI-RADS, alone.

**Results:** 151 patients met criteria for inclusion. AUC values for ADC, RSI_rs_, and PI-RADS were 0.50 [95% confidence interval: 0.41, 0.60], 0.76 [0.68, 0.84], and 0.78 [0.71, 0.85], respectively. RSI_rs_ (*p*=0.0002) and PI-RADS (*p*<0.0001) were superior to ADC for patient-level detection of csPCa. The performance of RSI_rs_ was comparable to that of PI-RADS (*p*=0.6). AUC for PI-RADS+RSI_rs_ was 0.84 [0.77, 0.90], superior to PI-RADS or RSI_rs_, alone (*p*=0.008, *p*=0.009).

**Conclusions:** RSI_rs_ was superior to conventional ADC and comparable to (routine, clinical) PI-RADS for patient-level detection of csPCa. The combination of PI-RADS and RSI_rs_ was superior to either alone. RSI_rs_ is a promising quantitative marker worthy of prospective study in the setting of csPCa detection.

**Disclosures:** MEH reports honoraria from Multimodal Imaging Services Corporation and research funding from General Electric Healthcare. AMD is a Founder of and holds equity in CorTechs Labs, Inc, and serves on its Scientific Advisory Board. He is a member of the Scientific Advisory Board of Human Longevity, Inc. and receives funding through research agreements with General Electric Healthcare. The terms of these arrangements have been reviewed and approved by the University of California San Diego in accordance with its conflict-of-interest policies. TMS reports honoraria from Multimodal Imaging Services Corporation, Varian Medical Systems, and WebMD; he has an equity interest in CorTechs Labs, Inc. and also serves on its Scientific Advisory Board. These companies might potentially benefit from the research results. The terms of this arrangement have been reviewed and approved by the University of California San Diego in accordance with its conflict-of-interest policies.

## Introduction

Worldwide, prostate cancer represents the second most common cancer diagnosis and the fifth leading cause of death in men^1^. Multiparametric magnetic resonance imaging (mpMRI) has been shown to improve detection of clinically significant cancer while reducing the detection of indolent tumors^2–6^. The standardized qualitative scoring system for mpMRI, Prostate Imaging Reporting & Data System (PI-RADS), has contributed to this early success and has been improved with updated versions 2 and 2.1^7,8^. However, concerns remain regarding variable interpretation of mpMRI across readers and institutions^9–12^ and the lack of a robust quantitative mpMRI metric^13^. Moreover, mpMRI and PI-RADS perform suboptimally for detection of cancers within the transition zone as compared to the peripheral zone^14,15^.

Diffusion-weighted imaging (DWI) is a key component of mpMRI. DWI is the most important determinant of lesion classification in the prostate’s peripheral zone, where the majority of prostate cancers arise. DWI is also important in the transition zone, though benign prostatic hyperplasia in the transition zone can often complicate interpretation. The standard quantitative DWI metric, conventional apparent diffusion coefficient (ADC), is inversely correlated with malignancies of various origins because of its relationship to hypercellularity^16^. However, ADC depends on multiple scan parameters, including b-value scheme, scanner used, echo time, pulse duration, and magnetic field, which can lead to variability in contrast and signal intensities. ADC is also influenced by extracellular effects that can confound the relationship with cellularity^17,18^. Taken together, these effects result in the poor reliability of ADC^13^. A more accurate and reliable quantitative diffusion MRI metric could improve detection of clinically significant prostate cancer.

Restriction spectrum imaging (RSI) is an advanced diffusion modeling framework that accounts for complex tissue microstructure by estimating contributions of distinct diffusion patterns in different tissue compartments^18^. A recent study found that male pelvic tissues can be optimally modeled with four diffusion compartments^19^. Prostate tumor conspicuity was highest using the model coefficient for the most restricted diffusion compartment, called here the RSI restriction score (RSI_rs_). Voxel-level prostate cancer detection was also more accurate using RSI_rs_ than ADC^20^.

Here, we evaluated RSI_rs_ as a quantitative marker for patient-level detection of higher-grade (grade group ≥2) prostate cancer in a larger, independent dataset. Performance was compared to ADC, the current quantitative diffusion marker, as well as to the qualitative PI-RADS scoring system (v2 and v2.1). We also considered the combination of expert-assigned PI-RADS categories with RSI_rs_. We hypothesized that, compared to conventional ADC, RSI_rs_ would have superior accuracy for detection of higher-grade prostate cancer on biopsy.

## Methods

### Study Population

This retrospective study included men who underwent RSI-MRI for suspected prostate cancer at UC San Diego between January 2017 and December 2019 and had a prostate biopsy within 180 days of MRI. RSI-MRI exams included standard mpMRI, as well as an RSI series with four *b*-values in a single acquisition. This study was approved by the UC San Diego Institutional Review Board.

### MRI Acquisition and Processing

Scans were collected on a 3T clinical MRI scanner (Discovery MR750, GE Healthcare, Waukesha, WI, USA) using a 32-channel phased-array body coil. The acquisition parameters are described in **Table 1**.

**Table 1.**
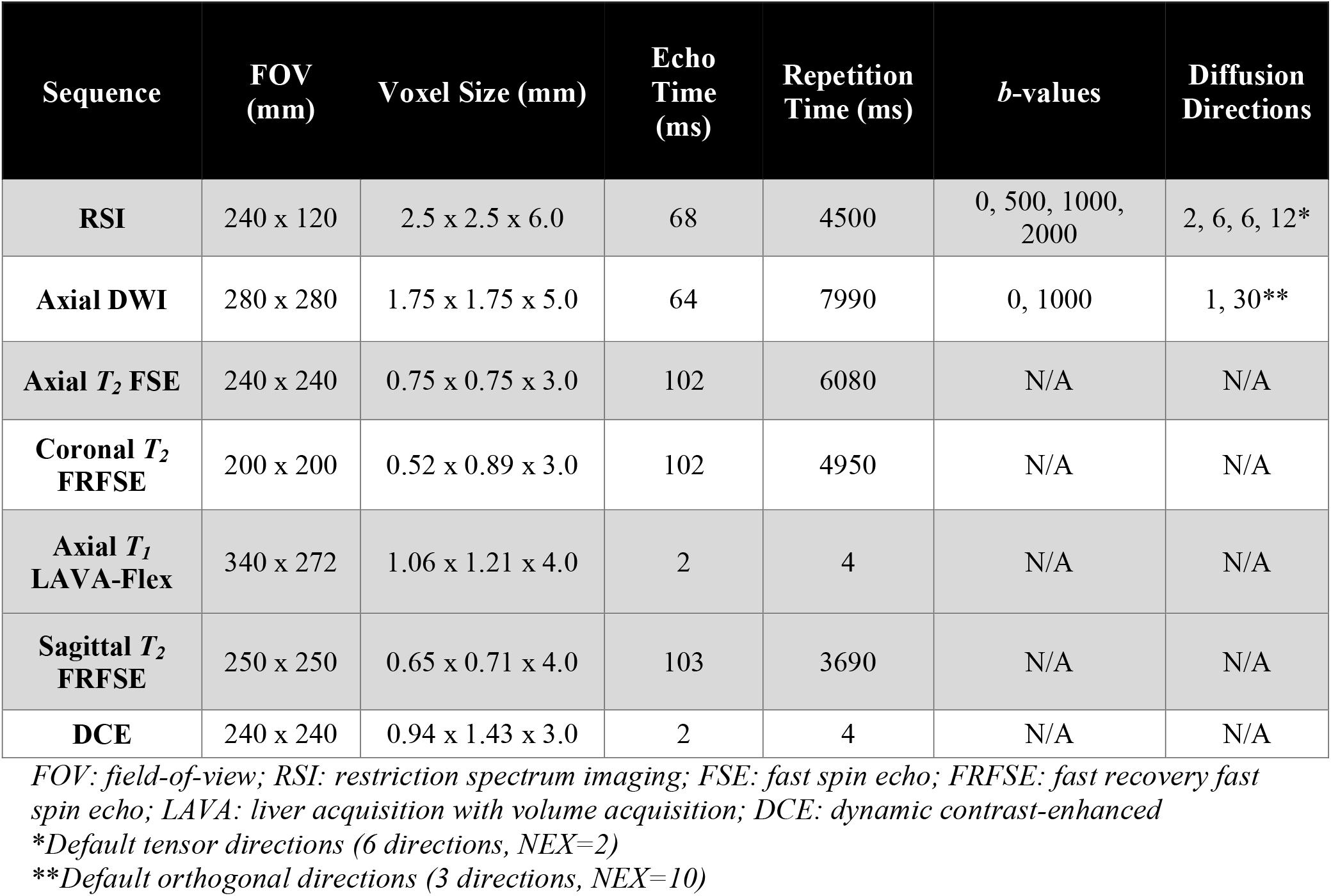
Acquisition parameters for clinical multiparametric (mp) MRI at 3T.

Post-processing of MRI data was completed using in-house programs written in MATLAB (MathWorks, Natick, MA, USA). Diffusion data were corrected for distortions arising from B0 inhomogeneity, gradient nonlinearity, and eddy currents^17,21^. RSI calculations were performed as previously described^19,20^. Briefly, diffusion signals were corrected for noise and distortion, then scaled by the median *b*=0 signal within each patient’s prostate. Corrected signal intensity for each *b*-value, *S*_*corr*_(*b*), was modeled as a linear combination of exponential decays representing four diffusion compartments (*i*=1-4) with fixed diffusion coefficients *D*_*i*_ and voxel-wise estimated signal contributions *C*_*i*_.

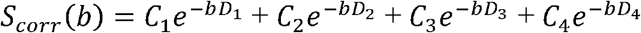

Optimal *D*_*i*_ values for each compartment were previously determined for pelvic and prostate tissues: 1.0 e-4, 1.8 e-3, 3.6 e-3, and >>3.0 e-3 mm^2^/s, approximately representing restricted, hindered, free diffusion, and flow, respectively^19^.

ADC maps were generated automatically, per clinical routine, using vendor software on the MRI system and the Axial DWI acquisition with *b*-values 0 and 1,000 s/mm^2^ (Table 1).

The prostate gland, peripheral zone, and central gland were manually segmented on T2-weighted imaging and verified on DWI volumes using MIM (MIM Software Inc, Cleveland, OH, USA).

### Clinical Data

Clinical records were reviewed to obtain histopathology results (highest grade group^22^ on biopsy or prostatectomy, if applicable) and imaging results (highest PI-RADS category reported). MRI exams were read by board-certified and subspecialty-fellowship-trained radiologists, using all available clinical images and standard PI-RADS criteria (transition to v2.1 from v2 occurred in 2019^7,8^).

### Statistical Analyses

#### Primary Analysis: Patient-level Detection of Higher-grade Prostate Cancer

Receiver operating characteristic (ROC) curves were generated for patient-level detection of higher-grade prostate cancer (grade group ≥2) using ADC, RSI_rs_, and PI-RADS. In the primary analysis, RSI_rs_ and ADC were analyzed as quantitative metrics, taking the maximum RSI_rs_ and minimum ADC within the prostate (analogous to the use of maximum standardized uptake value (SUV) in PET imaging for cancer^23^). Gleason ≤6 cancers or fully benign biopsies were considered negative results for the ROC curves. Performance was assessed by area under the ROC curve (AUC). Statistical comparisons were made via 10,000 bootstrap samples to calculate 95% confidence intervals and bootstrap *p*-values for the difference between the performance (AUC) of ADC, RSI_rs_, and PI-RADS^24^. Significance was set at two-sided α=0.05.

Using procedures analogous to those described above, subsequent analyses were conducted as follows:

#### Quantitative Diffusion MRI within PI-RADS Categories

To determine whether RSI_rs_ enhances the detection of higher-grade prostate cancer compared to PI-RADS alone, the patient-level analysis was repeated within the strata of each PI-RADS category (i.e., 2, 3, 4, and 5). As described above, the maximum RSI_rs_ and minimum ADC within the prostate were used. ROC curves were generated within each category, and within-PI-RADS-category performance was estimated via AUC.

#### Combination of PI-RADS and RSI

To evaluate overall performance of the combination of PI-RADS and RSI_rs_, an ROC curve for PI-RADS+RSI_rs_ was generated by fitting a logistic regression model within each PI-RADS category (for presence of higher-grade prostate cancer, using maximum RSI_rs_ within the prostate as the sole predictive variable) and concatenating the logistic regression posterior probabilities from the various PI-RADS strata. Overall performance of PI-RADS+RSI_rs_ was assessed by AUC of the resulting ROC curve. Performance of PI-RADS+RSI_rs_ was compared to either PI-RADS or RSI_rs_, alone, via the bootstrapping procedure described above.

#### Peripheral Zone and Central Gland

The patient-level analysis was repeated after categorizing cases by location within the prostate. Specifically, the analyses separately considered cases with lesions in the peripheral zone or central gland (including transition zone). For the peripheral zone analysis, patients diagnosed with a higher-grade cancer were excluded if they had a lesion detected on mpMRI in the transition zone (i.e., any PI-RADS category ≥2 assigned in the transition zone), and the search for the maximum RSI_rs_ and minimum ADC was limited to the peripheral zone. An analogous analysis was performed for the central gland, using PI-RADS lesions within the transition zone and limiting voxel-wise RSI_rs_ and ADC searches to only the central gland.

#### Impact of Artificial Errors in Prostate Segmentation

To assess the dependence of results on accurate prostate segmentation, the patient-level analysis was repeated after expanding whole prostate contours by a uniform 5-mm margin in all directions. The maximum RSI_rs_ and minimum ADC within the post-expansion segmentation were used. ROC curves and AUCs were calculated as before.

#### Alternate ADC Map

Vendor-calculated ADC maps were used for the above analyses in order to represent current clinical practice. However, as a secondary analysis, voxel-wise ADC was also calculated in MATLAB from the RSI acquisition, using b-values of 0, 500, and 1000 s/mm^2^. The above analyses were repeated, and the results using these alternate ADC maps were compared to the results using clinically standard, vendor-computed ADC maps.

## Results

151 patients met criteria for inclusion. The median age was 66 years (IQR 59-72). The median time from MRI to biopsy was 16 days (IQR 1-35). PI-RADS and pathology characteristics are summarized in **Table 2**. 104 patients were scored using PI-RADS v2, and 47 were scored using v2.1 after our institution transitioned to v2.1 in 2019.

**Table 2.**
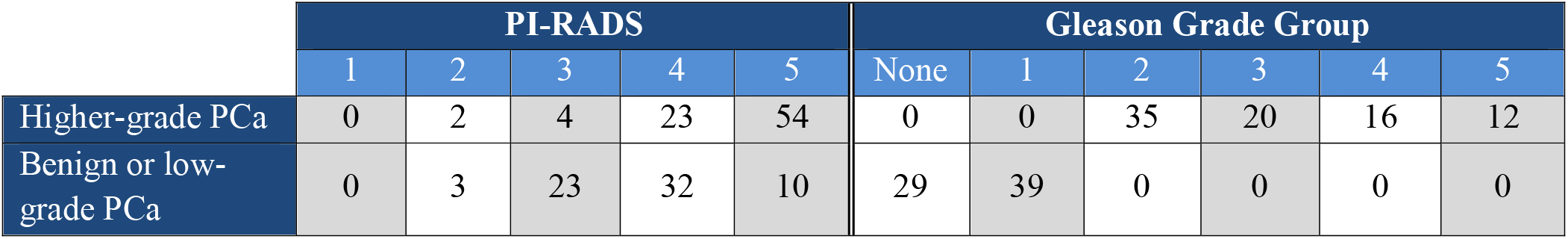
PI-RADS and pathology characteristics of patients included in this study.

### Primary Analysis: Patient-level Detection of Higher-grade Prostate Cancer

All 151 patients were included in the primary (whole-prostate) analysis. The AUC values for ADC, RSI_rs_, and PI-RADS were 0.50 [95% confidence interval: 0.41, 0.60], 0.76 [0.68, 0.84], and 0.78 [0.71, 0.85], respectively. Both RSI_rs_ (*p*=0.0002) and PI-RADS (*p*<0.0001) were superior to ADC as a patient-level classifier of higher-grade prostate cancer. The performance of RSI_rs_ was comparable to that of PI-RADS (*p*=0.6). The histograms and ROC curves for the primary analysis are shown in **Figure 1** and **Figure 2**, respectively.

**Figure 1.**
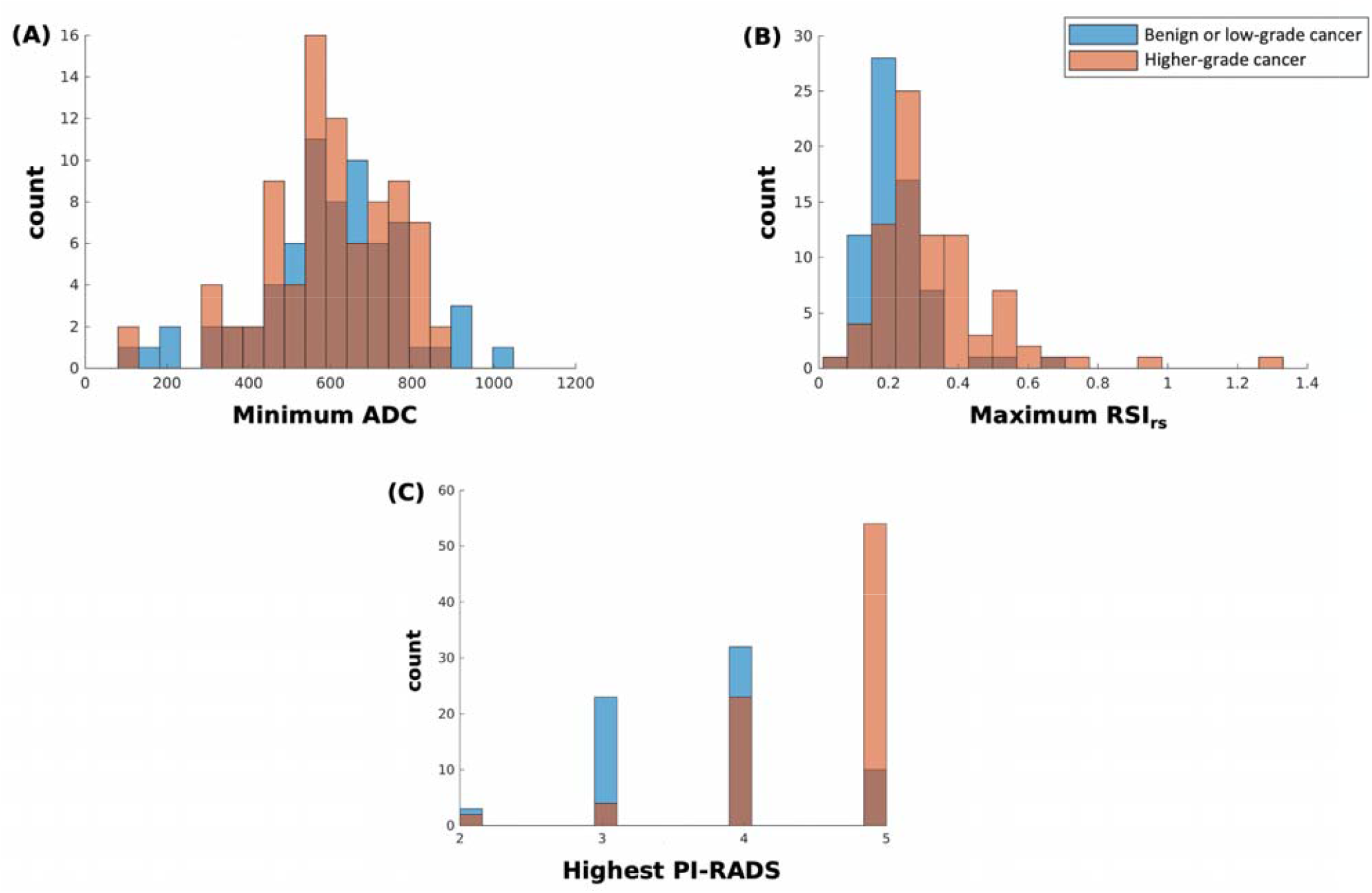
Histograms of (A) minimum conventional ADC in the prostate, (B) maximum RSI_rs_ in the prostate, and (C) highest PI-RADS category (v2 prior to 2019, v2.1 in 2019) in the prostate. Blue: Patients with no cancer or low-grade cancer. Orange: Patients with higher-grade (grade group ≥2) prostate cancer. Brown: Where blue and orange overlap.

**Figure 2.**
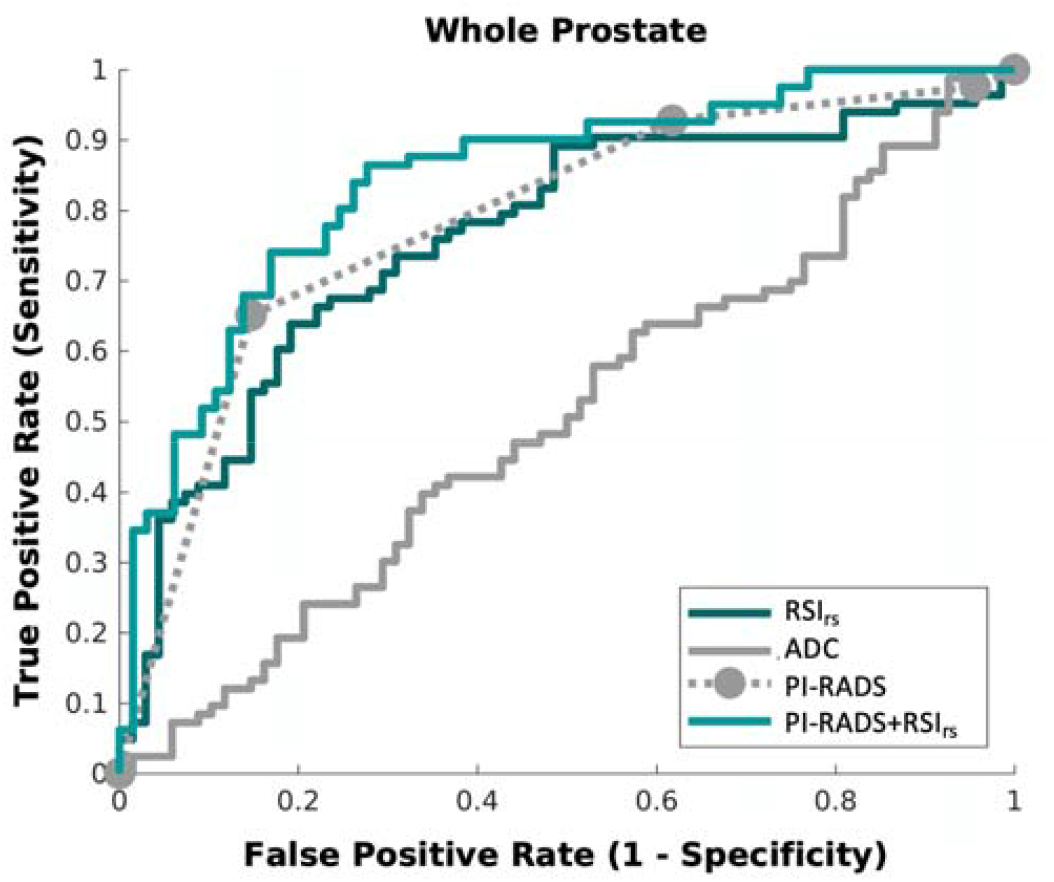
Receiving operator characteristic (ROC) curves for conventional ADC (solid gray), RSI_rs_ (dark green), PI-RADS (dashed gray), and PI-RADS+RSI_rs_ (light green) for patient-level detection of higher-grade prostate cancer anywhere in the prostate.

### Quantitative Diffusion MRI within PI-RADS Categories

There were 27, 55, and 64 patients included in PI-RADS groups 3, 4, and 5, respectively. The AUC values for maximum prostate RSI_rs_ within PI-RADS groups 3, 4, and 5 were 0.70 [0.50, 0.87], 0.69 [0.54, 0.83], and 0.70 [0.51, 0.86], respectively.

### Combination of PI-RADS and RSI

All 151 patients were included in the combination analysis. The AUC value for PI-RADS+RSI_rs_ was 0.84 [0.77, 0.90]. PI-RADS+RSI_rs_ was superior in performance to either PI-RADS (*p*= 0.008) or RSI_rs_ (*p*=0.009) alone (**Figure 2**).

### Peripheral Zone

103 patients without a transition zone lesion (15 benign, 23 grade group 1, 65 csPCa) were identified and included. For these peripheral zone cases, the AUC values for ADC, RSI_rs_, PI-RADS, and PI-RADS+RSI_rs_ were 0.50 [0.39, 0.62], 0.79 [0.69, 0.88], 0.80 [0.71, 0.87], and 0.89 [0.82, 0.95], respectively. The performance of RSI_rs_ was comparable to that of PI-RADS for the peripheral zone (*p*=0.9). ROC curves are shown in **Figure 3(a)**.

**Figure 3.**
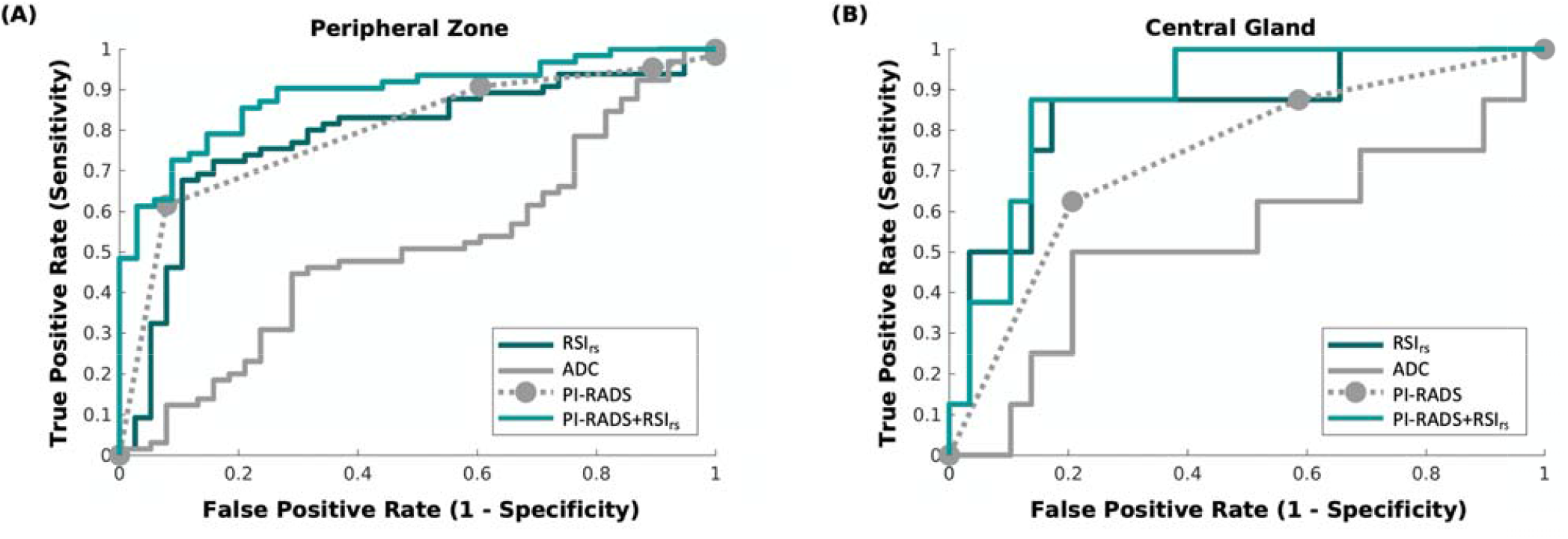
Receiving operator characteristic (ROC) curves for conventional ADC (solid gray), RSI_rs_ (dark green), PI-RADS (dashed gray), and PI-RADS+RSI_rs_ (light green) for patient-level detection of higher-grade prostate cancer (a) in the peripheral zone or (b) in the central gland.

### Central Gland

37 patients without a peripheral zone lesion (14 benign, 15 grade group 1, 8 csPCa) were identified and included. For these central gland cases, the AUC values for ADC, RSI_rs_, PI-RADS, and PI-RADS+RSI_rs_ were 0.53 [0.29, 0.78], 0.85 [0.67, 0.98], 0.74 [0.53, 0.90], and 0.89 [0.75, 0.98], respectively. There was a trend toward superior performance with RSI_rs_, compared to that of PI-RADS (*p*=0.05). ROC curves are shown in **Figure 3(b)**.

**Figure 4.**
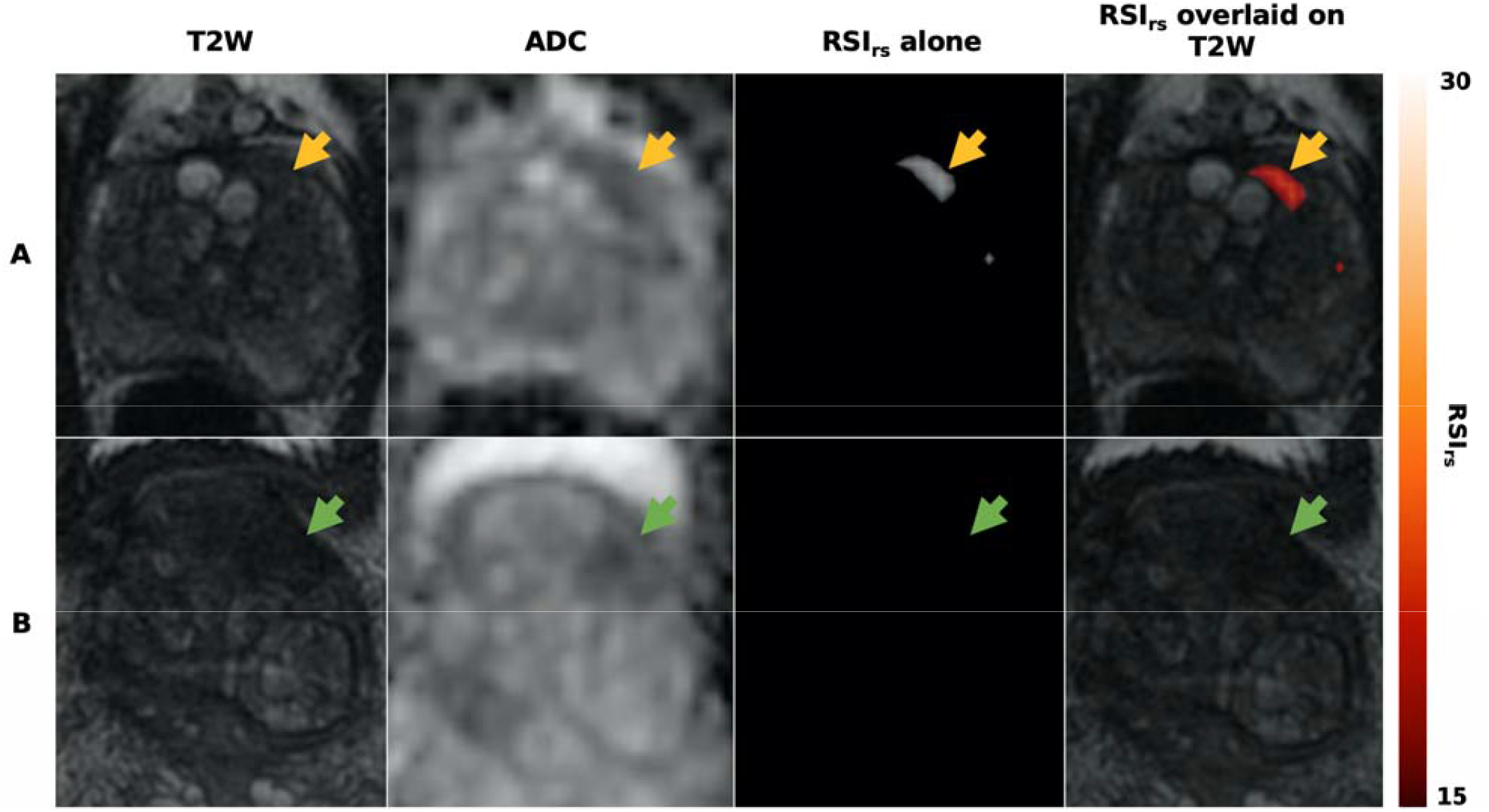
Axial images from two patients with transition zone lesions: *T*_*2*_-weighted MRI (T2W), conventional ADC, and RSI_rs_. Patient A had a PI-RADS 3 lesion (yellow arrow) in the left transition zone; he underwent prostatectomy and was found to have Gleason 3+4 prostate cancer. Patient B had a PI-RADS 5 lesion (green arrow) on multiparametric MRI, with subsequent biopsy showing benign prostatic tissue with acute and chronic inflammation. The RSI_rs_ map readily highlights the cancer for Patient A. The RSI_rs_ map for Patient B has no false-positive voxels (and is shown on the same color scale as the map for Patient A).

### Impact of Artificial Errors in Prostate Segmentation

All 151 patients were included in this secondary analysis. Prior to the expansion, the AUC values for ADC, RSI_rs_, and PI-RADS were 0.51 [0.41, 0.60], 0.76 [0.68, 0.84], and 0.78 [0.72, 0.85], respectively. Using the expanded region of interest, the AUC values for ADC, RSI_rs_, and PI-RADS were 0.51 [0.50, 0.61], 0.77 [0.69, 0.85], 0.78 [0.71, 0.85], respectively. The results did not meaningfully change after the expansion, suggesting maximum prostate RSI_rs_ is robust to inaccuracies in prostate segmentation that include peri-prostatic tissue.

### Alternate ADC Map

All 151 patients were included in this secondary analysis. The AUC value for the alternate ADC maps was 0.58 [0.49, 0.67], whereas that for the vendor-calculated ADC maps was 0.50 [0.41, 0.60]. There was no significant difference in performance (AUC) between the alternate ADC maps and vendor-calculated ADC maps (*p*=0.24).

## Discussion

Overall, RSI_rs_ performed well for quantitative, automated detection of higher-grade cancer within the prostate. In contrast, ADC, the current standard diffusion metric, proved unreliable as a quantitative marker for patient-level detection of csPCa with an analogous approach. RSI_rs_ was based solely on a two-minute diffusion MRI acquisition, yet performance was comparable to PI-RADS categories assigned by experts using all images (including ADC maps) from a complete multiparametric MRI exam. Moreover, combining PI-RADS categories and maximum RSI_rs_ improved performance over either alone.

Results here are consistent with prior work. A modeling study showed that the four-compartment RSI model used here yielded optimal explanation of diffusion signals within the pelvis and increased conspicuity of prostate cancer tumors^19^. Voxel-level accuracy of RSI_rs_ was superior to that of ADC^20^. Other work established the potential utility of RSI for cancer^17^, found voxel-wise correlation of RSI with Gleason pattern^18^, and better discrimination of PCa from normal tissue than ADC^25^. A retrospective analysis using a different RSI model did not find improvement over standard diffusion MRI for most cases but did show superior specificity for RSI in the transition zone^26^. In the present study, though relatively few csPCa cases could be included, performance for RSI_rs_ was also promising in the central gland, with a trend toward improvement over PI-RADS (*p*=0.05). The combination of PI-RADS and RSI_rs_ yielded excellent patient-level accuracy for csPCa in this dataset, with AUC of 0.88 [0.77, 0.98].

Moving toward quantitative imaging biomarkers is a stated aim of major imaging organizations and research funders, who have noted a paucity of validation studies^13,27^. The RSI approach adopted in the present work incorporates steps toward reproducibility, including distortion correction and normalization^20,28^. We have also demonstrated here the performance of a quantitative metric for cancer detection in a completely independent dataset from that used to develop the model. Additionally, the acquisition protocol used here is distinct from the data used to train the four-compartment RSI model^19^, including different *b*-values and echo time.

It is worth emphasizing that PI-RADS categories for this study were assigned during routine clinical practice. All readers were board-certified and sub-specialty trained attending radiologists at an academic center and adhered to PI-RADS standards, but this does not preclude some inter-reader variability. PI-RADS v2 was also replaced by PI-RADS v2.1 part-way through the time period studied. Nevertheless, overall performance shown here for PI-RADS is within the range of expected values^9^. Also of note, while the radiologists at our institution adhere to PI-RADS criteria for assigning lesion categories, they do have access to all images, including those of the RSI acquisition. Clinical decision-making surrounding biopsy may have been influenced by RSI images, in addition to PI-RADS, comments on possible extraprostatic extension, seminal vesicle invasion, or any number of non-imaging clinical factors.

Limitations include retrospective design, single institution, lack of prostatectomy specimen in all patients, and inclusion of only patients who underwent biopsy and had results available in our institutional patient records. This sample of convenience does not lend itself to evaluation of the true sensitivity of MRI for csPCa. However, it is not ethical in routine clinical practice to obtain biopsy (much less prostatectomy) specimens on all patients, and so all studies are subject to selection bias. Imaging for this dataset was all acquired on a single scanner; more reproducibility data are needed, including on other imaging systems. Still, the present study does represent an independent validation of RSI_rs_ in a new dataset obtained with a distinct imaging acquisition protocol.

In conclusion, this study demonstrates that RSI_rs_ achieved superior performance for patient-level detection of higher-grade prostate cancer than that of conventional ADC and comparable to that of routine, clinical PI-RADS. The combination of PI-RADS and RSI_rs_ performed better than either RSI_rs_ or PI-RADS, alone. Moreover, this pattern held true within the central gland, a region known to be more challenging for standard mpMRI. RSI_rs_ holds promise as a quantitative marker and should be prospectively studied for improvement of csPCa diagnosis.

## Data Availability

Contact the corresponding author with any inquiries regarding the data presented in this manuscript.

